# Disruption in seasonality, patient characteristics and disparities of respiratory syncytial virus infection among young children in the US during and before the COVID-19 pandemic: 2010-2022

**DOI:** 10.1101/2022.11.29.22282887

**Authors:** Lindsey Wang, Pamela B. Davis, Nathan A. Berger, David C. Kaelber, Nora D. Volkow, Rong Xu

**Affiliations:** Center for Science, Health, and Society, Case Western Reserve University School of Medicine, Cleveland, OH, USA; Center for Community Health Integration, Case Western Reserve University School of Medicine, Cleveland, OH, USA; The Center for Clinical Informatics Research and Education, The MetroHealth System, Cleveland, OH, USA; National Institute on Drug Abuse, National Institutes of Health, Bethesda, MD, USA; Center for Artificial Intelligence in Drug Discovery, Case Western Reserve University School of Medicine, Cleveland, OH, USA

## Abstract

Respiratory syncytial virus (RSV) infections and hospitalization have surged sharply among young children. Here we test how the seasonal patterns of RSV infections in 2022 compared with those from other COVID-19 pandemic and pre-pandemic years. For this purpose, we analyzed a nation-wide and real-time database of electronic health records of 56 million patients across 50 states in the US. The monthly incidence rate of first-time RSV infection in young children (<5 years of age) and very young children (<1 year of age) followed a seasonal pattern from 2010 to 2019 with increases during the autumn, peaking in winter, subsiding in spring and summer. This seasonal pattern was significantly disrupted during the COVID-19 pandemic. In 2020, the incidence rate of RSV infections was remarkably low throughout the year. In 2021, the RSV season expanded to 9 months starting in the early summer and peaking in October. In 2022, RSV infections started to rise in May and were significantly higher than in previous years reaching a historically highest incidence rate in November 2022. There were significant racial and ethnic disparities in the peak RSV infection rate during 2010-2021 and the disparities further exacerbated in 2022 with peak incidence rate in black and Hispanic children 2-3 times that in white children. Among RSV-infected children in 2022, 19.2% had prior documented COVID-19 infection, significantly higher than the 9.7% among uninfected children, suggesting that prior COVID-19 could be a risk factor for RSV infection or that there are common risk factors for both viral infections. Our study calls for continuous monitoring of RSV infection in young children alongside its clinical outcomes and for future work to assess potential COVID-19 related risk factors.

## Introduction

Respiratory syncytial virus (RSV) is a leading cause of lower respiratory tract infection in young children^1^. The COVID pandemic disrupted the circulation of RSV and other respiratory viral infections in the United States for 2020-2021^2^. In 2022 high rates of hospitalization with RSV infections have been reported particularly among the youngest children^3,4^. However, the magnitude, timing, and duration of this surge in RSV infections in young children remains unknown. Here we assessed the seasonal pattern of first-time RSV infection among young children in the US in 2022 and compared it to other pandemic and pre-pandemic years. For this purpose, we leveraged a nation-wide and real-time database of electronic health records (EHRs) of 56 million patients in the US and examined seasonal patterns and patient characteristics of RSV infections among young children ≤ 5 years of age and very young children ≤ 1 year of age from 2010 through November 2022. We compared characteristics of RSV-infected children with uninfected children in order to identify potential COVID-19 related risk factors.

## METHODS

### Study population

We used the TriNetX Analytics Platform to access aggregated and de-identified electronic health records (EHRs) of 56 million patients from 34 health care organizations in the US across 50 states, covering diverse geographic, age, race/ethnic, income and insurance groups.^5^ TriNetX built-in analytic functions allow for patient-level analyses, while only reporting population level data. The MetroHealth System, Cleveland OH, IRB determined research using TriNetX is not Human Subject Research and therefore IRB exempt. We previously used TriNetX Analytics network platform to study risk factors and outcomes of COVID-19 and other diseases^6–16^, including examining the incidence rates and clinical outcomes of COVID-19 with the Omicron and Delta variants in children ≤ 5 years of age^6^.

We examined the monthly incidence rate of first-time diagnosis of RSV infection (measured by new cases per 1,000,000 person-days) from 2010 through November 2022, among children ≤ 5 years of age and children ≤ 1 year of age who had a medical visit with a healthcare organization and had no prior RSV infection. The same analyses were performed in children ≤ 5 years of age stratified by race and ethnicity (Black, White and Hispanic), and by gender (Female and Male). We compared characteristics of children who were infected with RSV for the first-time during May-November 2022 to uninfected children during the same time frame. The characteristics that were compared include demographics, prior COVID-19 infection, RSV-related comorbidities^17^, adverse socio-economic determinants of health including physical, social and psychosocial environment, and housing, and COVID-19 vaccination status. Separate analyses were performed for children ≤ 1 year of age. All the analyses were conducted on 11/24/2022 within the TriNetX Analytics Platform with significance set at p-value < 0.05 (two-sided).

## Results

### Seasonal pattern of monthly incidence rate of first-time RSV infection among young children in the US during 2010-2022

We examined electronic health records of 42,222,538 medical visits for children ≤ 5 years of age and 12,507,431 medical visits among children ≤ 1 year of age from 2010 to 2022 (study ending date of November 23, 2022). From 2010 through 2019, the monthly incidence rate of RSV infection in children ≤ 5 years of age followed a consistent seasonal pattern: starting to rise from September to November, peaking from December to January, then dropping from February to April, with sustained decrease during May-August (**Figure 1a**). There was a significant rise in peak incidence rate in 2019 compared with previous years (561 cases per 1,000,000 person-days in December 2019 vs 335 cases per 1,000,000 person-days in December 2018, p<0.001). In 2019, there were higher cases in the summer months than for prior years. These data suggest the possible emergence of a new RSV strain in 2019 with higher infection rate and early seasonal start. Indeed, in earlier years, breakout infection with RSV during the summer in Minnesota was associated with a new strain of RSV-B^18^.

**Figure 1.**
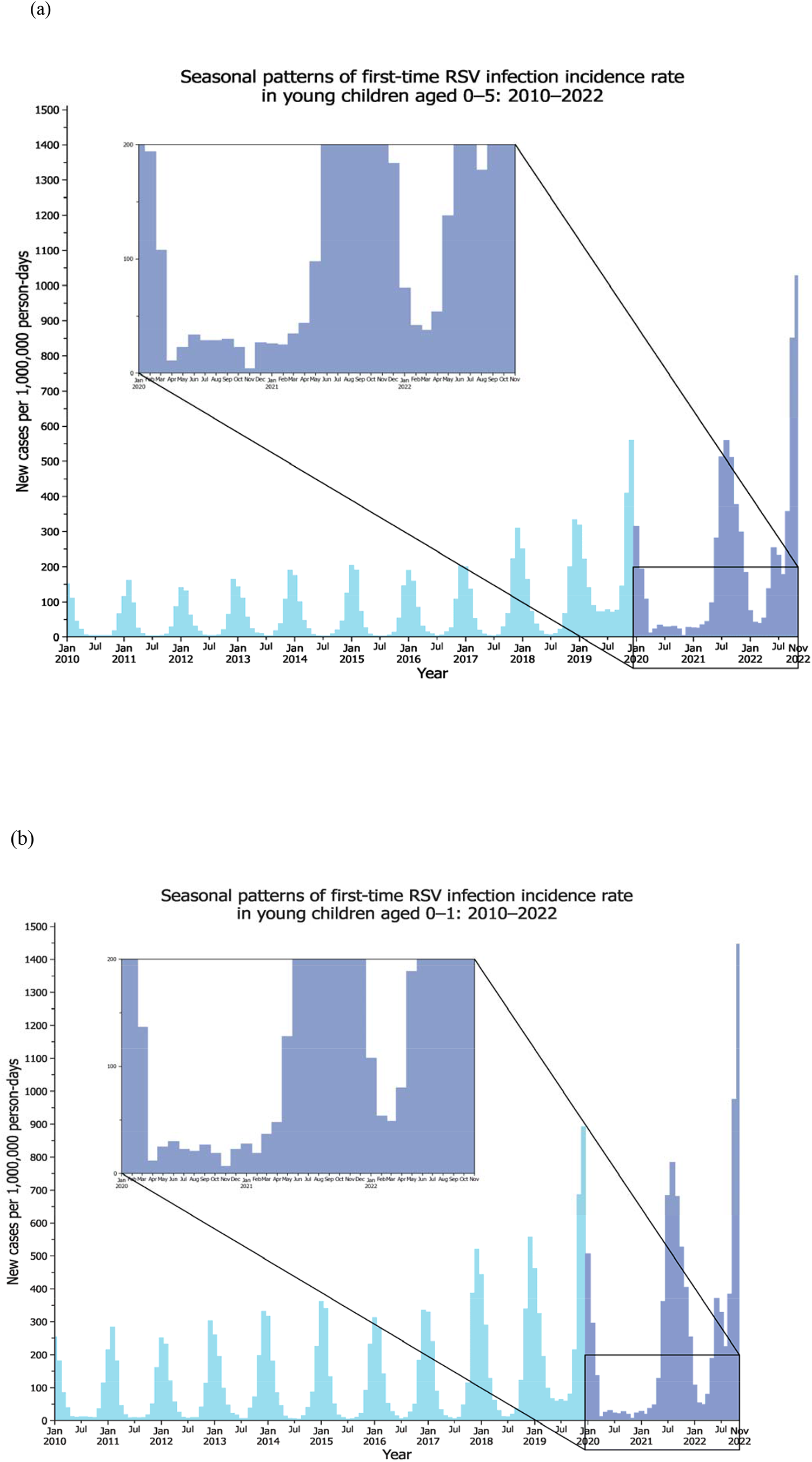
(a) Time trend of monthly incidence rate of RSV infection among children ≤ 5 years of old in the US during 2010-2022 with ending date of November 2022. (b) Time trend of monthly incidence rate of RSV infection among children ≤ 1 year of age in the US during 2010-2022 with ending date of November 2022. Monthly incidence rates were calculated as number of cases per 1,000, 000 person-days. The insert figures highlight the magnified pandemic years.

During the COVID-19 pandemic, the seasonal pattern of RSV infection changed. In 2020, seasonal variation disappeared, and the incidence rate was consistently low from May 2020 through April 2021 (ranging from 4 to 44 cases per 1000000 person-days) without a rise in the autumn or a peak in winter months. In 2021, the seasonality returned but started earlier than in pre-pandemic years with incidence rates starting to rise in May and June (98 and 282 cases per 1000000 person-days respectively), peaking in August (559 cases per 1,000,000 person-days) and starting to drop in October (377 cases per 1,000,000 person-days). The RSV season in 2021 expanded to 9 months (May 2021 – January 2022), longer than that in pre-pandemic years. The peak incidence rate in 2021 was similar to that in 2019, both significantly higher than in 2010-2018.

In 2022, the seasonal pattern changed again: the sustained rise continued from May (138 cases per 1,000,000 person-days) and reached a historically high incidence rate in November (1,027 cases per 1000000 person-days) (**Figure 1a**). Furthermore, the incidence rate was higher in 2022 than in any other previous years including for peak months in 2021. For example, the incidence rate among 86,878 children ≤ 5 years old in November 2022 was 1,027, higher than the peak incidence rate of 559 in August 2021.

Incidence rates of RSV infections in children ≤ 1 year old follow the same seasonal pattern as for children ≤ 5 years old, but with even higher rates. In November 2022, the incidence rate among 39,812 children ≤ 1 year of age was the highest ever recorded corresponding to 1,447 cases per 1,000,000 person-days (**Figure 1b**).

### Seasonal pattern of first-time RSV infection among young children by race, ethnicity, and gender

There were racial and ethnic differences in peak incidence rates of RSV infections among children ≤ 5 years of old. During 2010-2021, the incidence rates were significantly higher in Blacks than in Whites ranging from 8.9% to 44.0% (p<0.001), except in 2011 and 2017 when the rates were similar. In 2022, the difference in peak incidence rate further widened to 195% (2,135 cases per 1,000, 000 person-days in black children vs 723 cases per 1,000,000 person-days in white children) in November 2022 (**Figure 2a & 2b**). Higher peak incidence rates of RSV infections were observed in Hispanic children compared to white children, and the difference was larger than for Black vs White children. During 2010-2021, the incidence rates were consistently higher in Hispanic than in white children ranging from 7.0% to 50.6% (p<0.001). In 2022, the difference increased to 278.1% with 2,734 cases per 1,000, 000 person-days in Hispanic children in November 2022 (**Figure 2a & 2c**).

**Figure 2.**
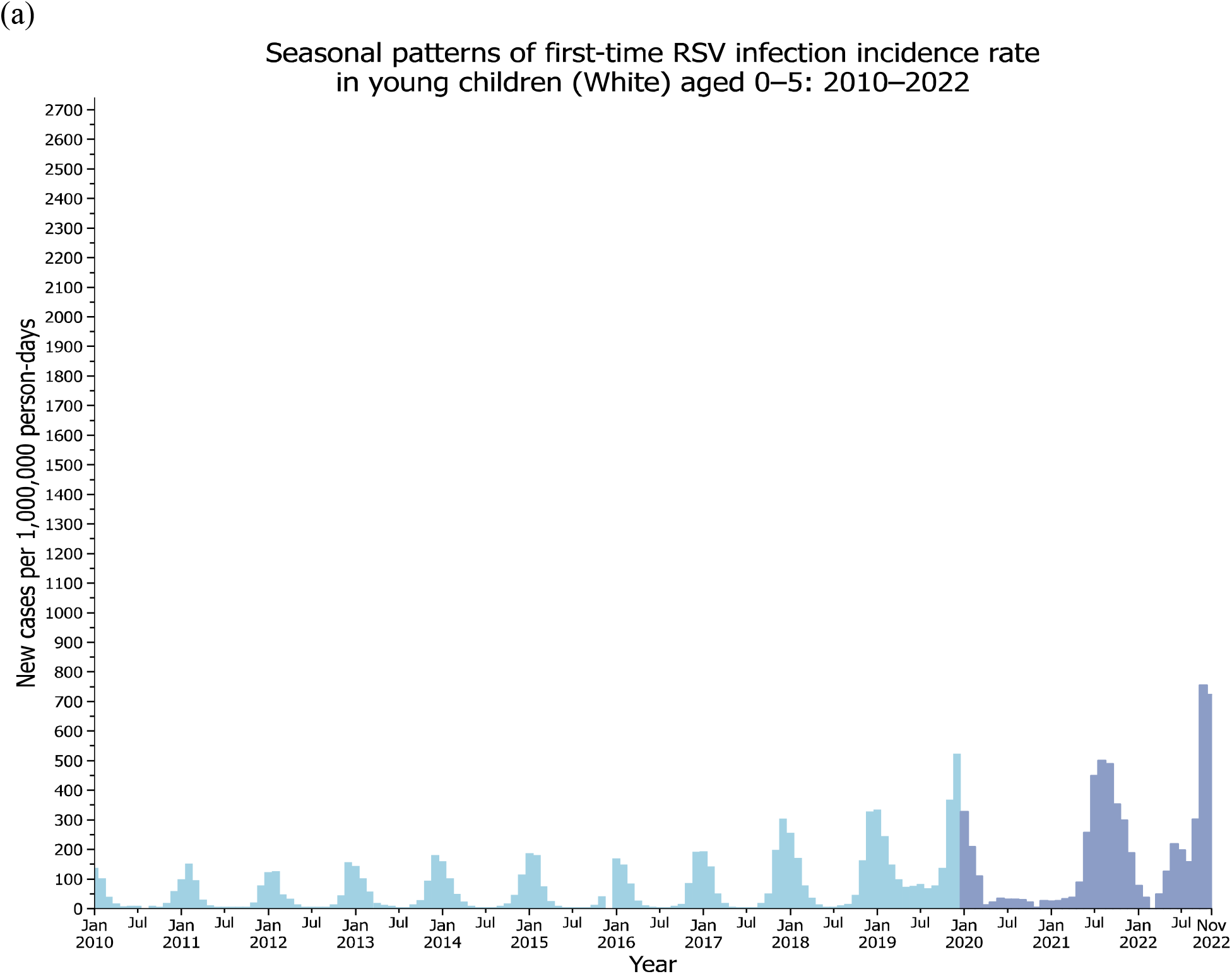
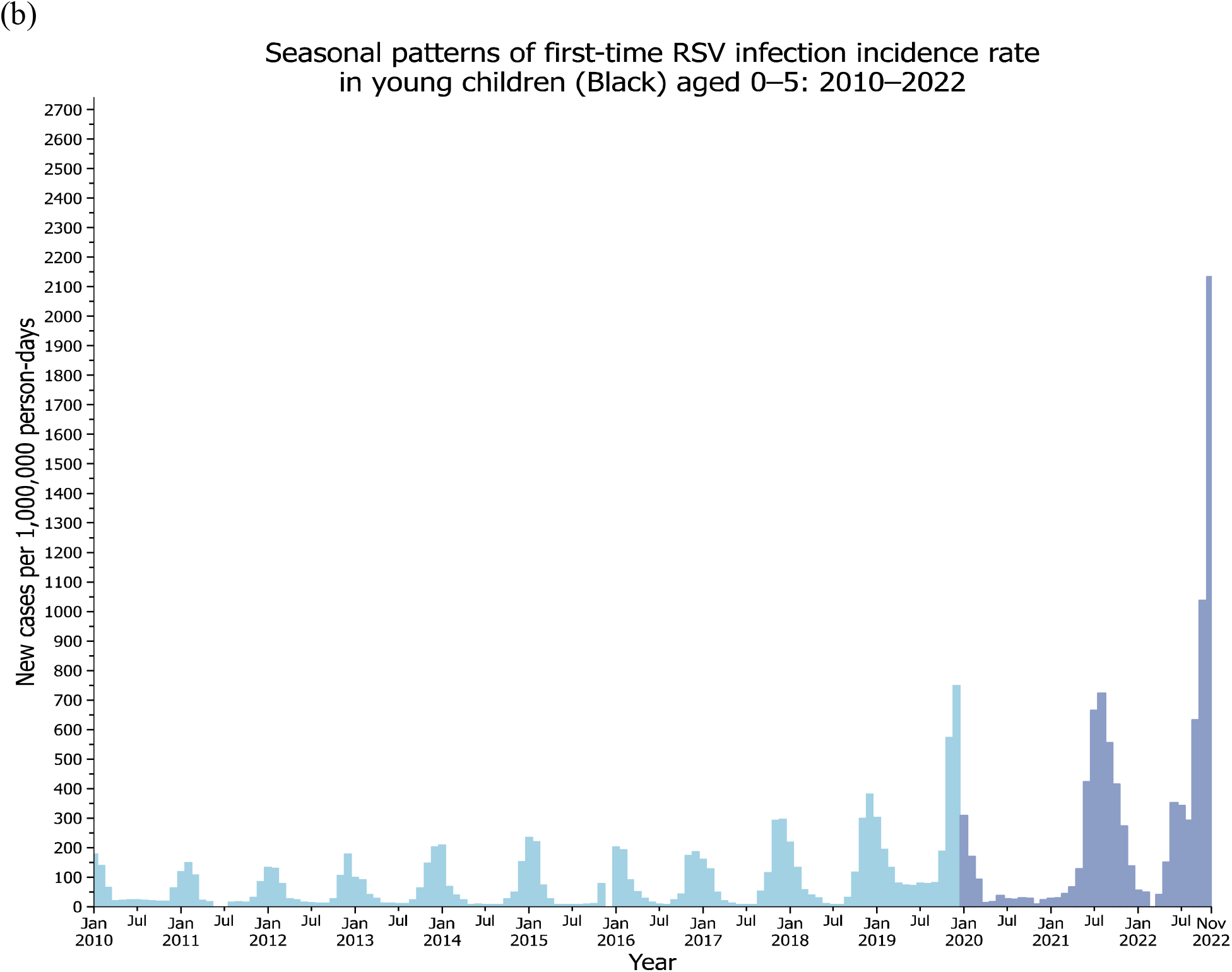
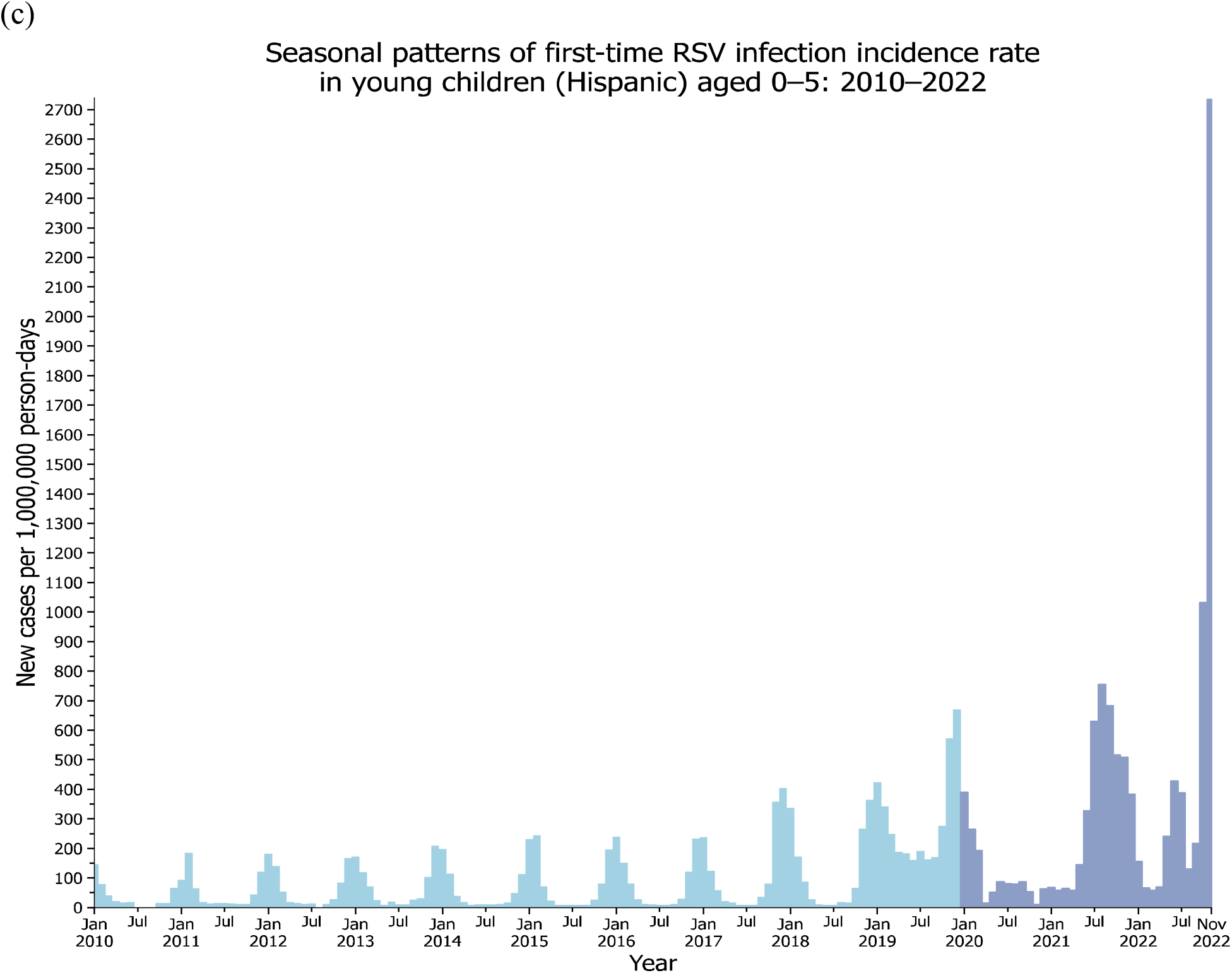
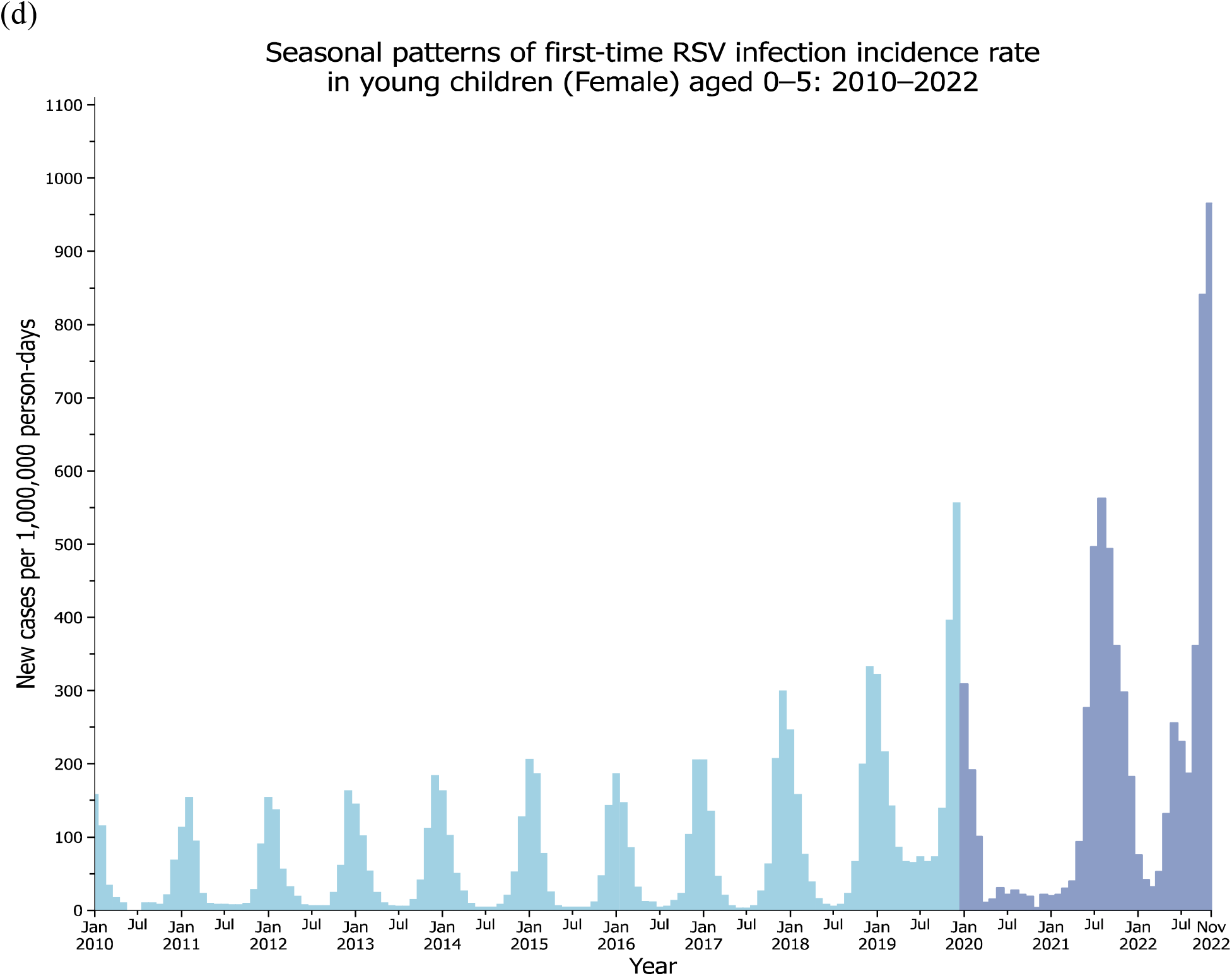
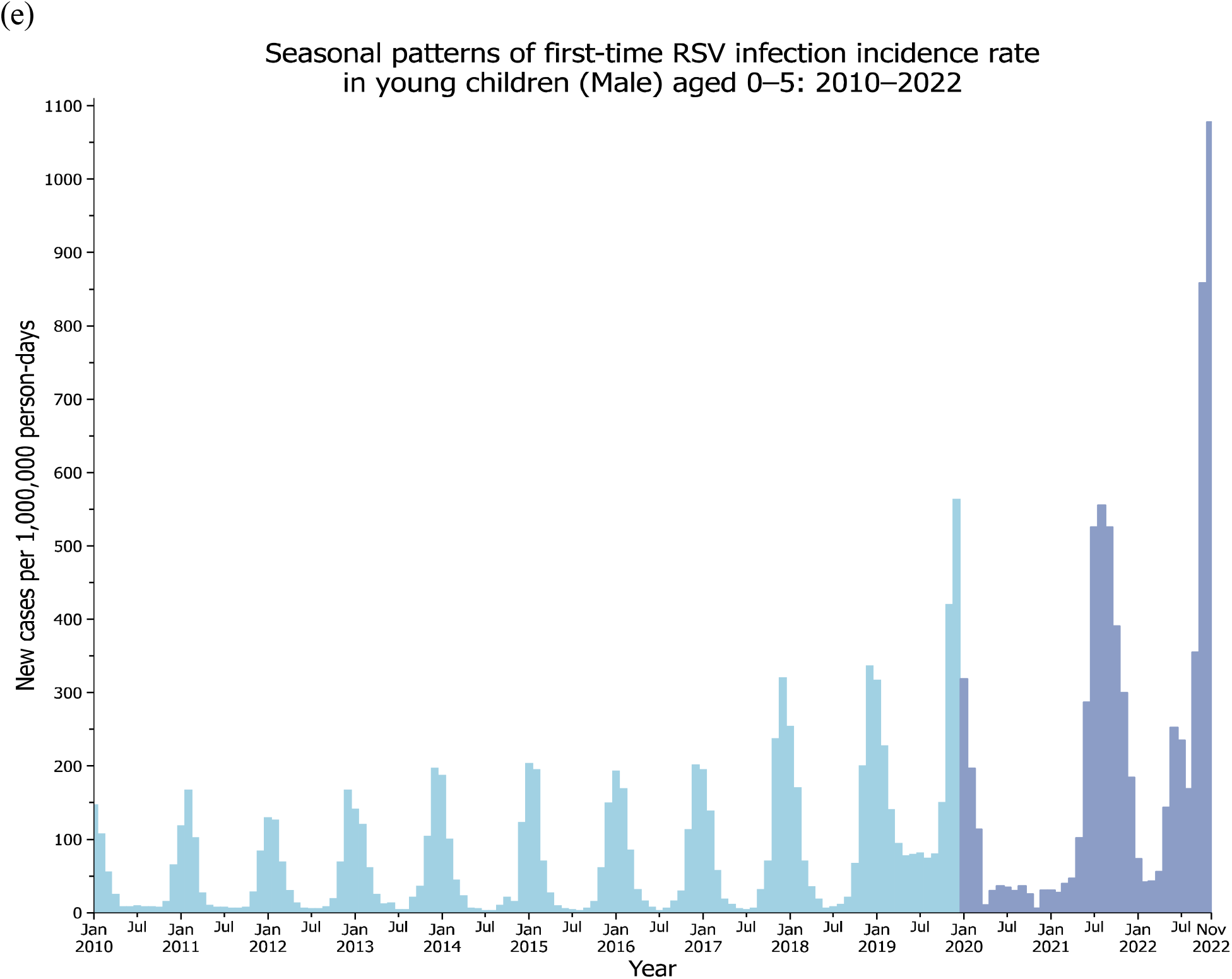
Time trend of monthly incidence rate of first-time RSV infections among children ≤ 5 years of old in the US during 2010-2022 stratified by race, ethnicity, and gender: (a) Black. (b) Hispanic, (c) White, (d) Female, and (d) male. Monthly incidence rates were calculated as number of cases per 1,000, 000 person-days.

There were no significant differences in peak incidence rates of RSV infections during 2010-2021 between boys and girls, with difference fluctuating between -19.2% to 7.7%. However in 2022, boys had a significantly higher peak incidence rate than girls in November 2022 (1,078 vs 966 cases per 1,000, 000 person-days) (p<0.001) (**Figure 2d & 2e**).

### Comparison of characteristics of RSV-infection children with uninfected children in 2022

We compared characteristics of young children who contracted RSV for the first-time during May-November 2022 (RSV (+) cohort) to those who did not (RSV (-) cohort) during the same time-period. Among 601,293 children ≤ 5 years of old who had no prior RSV and had a medical encounter with a healthcare organization during May-November 2022, 7,823 contracted RSV. Compared to the RSV (-) cohort, RSV (+) cohort was significantly younger, comprised more Hispanics and Blacks, had higher prevalence of adverse socio-economic determinants of health and higher comorbidities that are known risk factors for RSV infection including preterm birth, immune related disorders, chronic respiratory disease originating in the perinatal period, Downs syndrome and malnutrition. Furthermore, the RSV (+) cohort had higher prevalence of a prior COVID-19 infection than the RSV (-) cohort (19.2% vs 9.7%), suggesting the existence of common risk factors for both SARS-CoV-2 and RSV viral infections or the possibility that COVID-19 could constitute a potential risk factor for RSV among young children. No difference was found for COVID-19 vaccination status (**Table 1**). Similar findings were observed for children ≤ 1 year old.

**Table 1.** Characteristics of young children with and without RSV infection. Age was based on current age as of November 23, 2022. The status for adverse socioeconomic determinants of health (including physical, social and psychosocial environment, and housing) and RSV-related predisposing medical conditions were based on presences of related ICD-10 diagnosis codes in patient EHRs anytime up to November 23, 2022. The status of prior COVID-19 infection was based on EHR-documented presence of COVID-19 diagnosis code (ICD-10 diagnosis code U07.1) or positive COVID-19 lab test results at any time before RSV infection. The COVID-19 vaccination status was based on the presence of COVID-19 vaccination codes in patient EHRs anytime up to November 23, 2022.

|  | **2022 (May-November)**<br>**(Children aged $\leq 5$ years of age)** |  |  | **2022 (May-November)**<br>**(Children aged $\leq 1$ year of age)** |  |  |
| --- | --- | --- | --- | --- | --- | --- |
|  | **RSV (+)** | **RSV (-)** | **P-value** | **RSV (+)** | **RSV (-)** | **P-value** |
| **Total number** | 7,823 | 593,470 |  | 4,372 | 238,033 |  |
| **Current age (as of 11/23/2022) (years, mean±SD)** | 1.90 ± 1.66 | 2.59 ± 1.81 | <.001 | 0.69 ± 0.69 | 0.78 ± 0.69 | <.001 |
| **Sex (%)** |  |  |  |  |  |  |
| Female | 46.2 | 46.9 | 0.20 | 44.3 | 46.9 | <.001 |
| Male | 53.8 | 53.1 | 0.20 | 55.7 | 53.1 | <.001 |
| **Ethnicity (%)** |  |  |  |  |  |  |
| Hispanic/Latinx | 21.7 | 15.0 | <.001 | 21.8 | 15.8 | <.001 |
| Not Hispanic/Latinx | 62.8 | 57.6 | <.001 | 63.1 | 55.2 | <.001 |
| Unknown | 15.5 | 27.4 | <.001 | 15.1 | 29.0 | <.001 |
| **Race (%)** |  |  |  |  |  |  |
| Asian | 3.3 | 2.9 | 0.04 | 2.7 | 2.8 | 0.69 |
| Black | 20.1 | 17.1 | <.001 | 19.6 | 17.1 | <.001 |
| White | 54.8 | 58.2 | <.001 | 55.3 | 56.2 | 0.24 |
| Unknown | 21.3 | 21.2 | 0.77 | 22.0 | 23.3 | 0.47 |
| **Adverse socioeconomic determinants of health (%)** | 6.2 | 4.7 | <.001 | 4.8 | 3.2 | <.001 |
| **Pre-existing medical conditions (%)** |  |  |  |  |  |  |
| COVID-19 | 19.2 | 9.7 | <.001 | 17.2 | 9.2 | <.001 |
| Preterm birth, low birth weight | 6.0 | 5.0 | <.001 | 5.6 | 5.2 | 0.29 |
| Immune related disorders | 11.9 | 8.4 | <.001 | 8.3 | 5.1 | <.001 |
| Chronic respiratory disease originating in the perinatal period | 1.4 | 0.7 | <.001 | 1.1 | 0.8 | 0.08 |
| Congenital heart diseases | 4.9 | 4.6 | 0.35 | 4.2 | 4.5 | 0.38 |
| Neuromuscular Disorders | 0.4 | 0.4 | 0.65 | 0.2 | 0.2 | 0.36 |
| Downs syndrome | 0.6 | 0.3 | <.001 | 0.6 | 0.3 | <.001 |
| Malnutrition | 1.0 | 0.8 | 0.03 | 0.8 | 0.6 | 0.10 |
| **COVID-19 vaccine documented in EHRs (%)** | 3.5 | 3.5 | 0.84 | 2.2 | 2.4 | 0.53 |

## Discussion

To our best knowledge, our study is the first to examine nation-wide temporal seasonal patterns of first-time RSV infections in 2010-2022 in young and very young children that were further stratified by race, ethnicity and gender.

The monthly incidence rate of RSV infection in young children in the US during 2010-2018 followed the same seasonal pattern reported by CDC based on surveillance data from 2014-2017 ^19^; corroborating its validity. In 2019, there was a significant rise in peak incidence rate compared with previous years. Furthermore, there were higher cases in the summer months than for prior years. These data show a trend toward a change in the temporal pattern prior to the COVID-19 pandemic, suggesting the possibility of the emergence of a new RSV strain in 2019 with higher infection rate and early seasonal start.

The seasonal pattern of RSV infections was disrupted during the COVID-19 pandemic. In 2020, the incidence rate of RSV infections was low throughout the year, which may be due to lock-down, school and daycare closure, and COVID mitigation measures such as masking and social distancing, all contributing to reduce RSV exposure. In 2021 in contrast to 2020, the RSV season expanded to 9 months: the incidence rate of RSV began to rise in the summer months, peaked in the autumn and started to decline in the winter. This shift of RSV season to earlier months was also reported by the CDC surveillance data from 2020 to May 2021^2^. This may correspond to relaxation of COVID-19 mitigation strategies Findings from our study further showed that the 2021 RSV season expanded to 9 months and peaked in August with a similar peak incidence rate as in 2019.

In 2022, this shift of RSV season to earlier months was also observed. In addition, the incidence rate of RSV infection was significantly higher than in 2021 and in pre-pandemic years and reached an historically high rate in November 2022 (the last date of this study), and the peak is still not established. In 2022, schools and nurseries were open and preventative measure and masking were largely relaxed. However, the infection pattern of RSV remains different from that prior to 2019 and importantly significantly higher, suggesting the possibility that COVID-19 or COVID-related risk factors including lasting impact on immune systems that may partially account for high RSV incidence rates in young children.

Our results revealed significant racial and ethnic disparities in peak RSV infection rate with higher rates in Black and Hispanic than in White children in 2010-2021. The disparities were further exacerbated in 2022 when the peak incidence rate in Black and Hispanic children was 2-3 times of that in White children. Socioeconomics factors such as crowded living conditions might contribute to the racial and ethnic disparities in RSV infections. However reasons underlying the markedly exacerbated disparities in 2022 compared with 2021 and pre-pandemic years warrant further investigation. No consistent gender differences were observed in the seasonal patterns of RSV infections in 2010-2022.

We showed that children who contracted RSV were younger, more often Black and Hispanic, and had significantly higher prevalence of known risk factors for RSV infection^17^, some of which are also risk factors for COVID-19 infection, including those associated with social and demographic factors including housing, physical, social environment and racial/ethnic minority status^20^. Indeed, in our study the rate of prior COVID-19 as documented in EHRs among RSV infected children was twice that in uninfected children, suggesting that COVID-19 consequences might also be a risk factor for RSV infection. This potential association requires further study, with control for potential confounders including temperature and geographic location.

Our study has several limitations: First, potential biases could be introduced by the observational and retrospective nature of analyses of EHRs. Second, there may be over/mis/under-diagnosis of RSV infections and other diseases in patient EHRs. However, the data were drawn from the same TriNetX dataset, therefore these issues should not substantially affect the comparative seasonal pattern analysis of RSV infections. Third, patients in the TriNetX database are those who had medical encounters with healthcare systems contributing to the TriNetX Platform.

Therefore, they do not necessarily represent the entire US population and results from the TriNetX platform need to be validated in other populations. Fourth, the EHRs data were drawn from 34 health care organizations in the US across 50 states, covering diverse geographic regions including 44% in Northeast, 23% in Midwest, 22% in South, and 10% in West. However, we could not further break down the trend patterns by regions due to the de-identification restrictions in TriNetX. Nonetheless, all the data from 2010-2022 were from the same 34 healthcare organizations, which should not significantly affect the comparative analysis of seasonal patterns of RSV infections. Despite the above mentioned limitations, our study confirmed the same seasonal pattern reported by CDC based on surveillance data from 2014-2017 ^19^, low RSV infection rate in 2020 and early seasonal start of RSV activity in 2021 consistent with CDC’s reported surveillance data from 2020 to May 2021^2^. These corroboration results demonstrate the validity of our EHR-based study for real-time monitoring RSV infection patterns in real-world patients. Fifth, many children have contracted COVID-19 though the actual prevalence is unknown^21^. The status of prior COVID-19 in our study was determined based on the clinical diagnosis code or positive lab test results captured in patients’ EHRs, which very likely was an under-estimate of the actual rate, because many COVID-19 tests were performed at home. However, we observed a two-fold difference in prior COVID-19 infection between RSV (+) and RSV (-) cohorts, suggesting that COVID-19 itself or common associated risk factors might contribute to the increased RSV infections among young children in 2022.

Sixth, our analyses cannot distinguish if the increased rates in RSV infection from the EHR reflect a rise in overall infections or in the severity of their presentation that led to seeking healthcare attention. Further research is needed to compare the clinical outcomes of RSV infections in 2022 with those in other pandemic and pre-pandemic years. Finally, social determinants of health (SDoH) are important to understanding patient health. The EHR data that we used captured substantial information on SDoH of the study population. As shown in Table 1, the percentage of the study population with the ICD-10 codes Z55-Z65 (“Persons with potential health hazards related to socioeconomic and psychosocial circumstances”) was 6.2% and 4.7% for RSV (+) and RSV (-) cohorts among young children. We observed significant difference between the two cohorts, but it remains unknown how complete and accurate these EHR-derived structured data elements capture SDoH.

In summary, we show that the seasonal pattern of RSV in young children was disrupted during the pandemic. The incidence rate of RSV in 2022 was significantly higher than that in previous years. The rate of prior COVID-19 infection for children who contracted RSV was double that from children who did not contract RSV. There were significant racial and ethnic disparities in peak RSV infection rate during 2010-2021, with the disparities further exacerbated in 2022. Our study by analyzing a nation-wide and real-time database of patient EHRs establishes an effective way to monitoring infection patterns of RSV in young children. The same data and methods can be readily applied to other infectious diseases and patient populations. Further research is warranted to examine the severity of RSV infection in 2022 compared with those in previous years in young children as well as in other age groups including older adults.

## Data Availability

All data produced in the present work are contained in the manuscript

## Contributors

RX conceived, designed, and supervised the study and drafted the manuscript. LW performed data analysis and prepared tables and figures and participated in manuscript preparation. LW, NDV, NAB, PBD, DCK critically contributed to study design, result interpretation and manuscript preparation. We confirm the originality of content. RX had full access to all the data in the study and takes responsibility for the integrity of the data and the accuracy of the data analysis.

## Acknowledgments

We acknowledge support from National Institute on Aging (grants nos. AG057557, AG061388, AG062272), National Institute on Alcohol Abuse and Alcoholism (grant no. R01AA029831), National Institute on Drug Abuse (grant no. UG1DA049435), the Clinical and Translational Science Collaborative (CTSC) of Cleveland (grant no. 1UL1TR002548-01), National Cancer Institute Case Comprehensive Cancer Center (R25 CA221718, P30 CA043703). R.X had full access to all the data in the study and takes responsibility for the integrity of the data and the accuracy of the data analysis.

## Meeting Presentation

No

